# Associations between D_3_Cr muscle mass and MR thigh muscle volume with strength, power, physical performance, fitness, and limitations in older adults in the SOMMA study

**DOI:** 10.1101/2023.11.09.23298303

**Authors:** Peggy M. Cawthon, Terri L. Blackwell, Stephen B. Kritchevsky, Anne B. Newman, Russell T. Hepple, Paul M. Coen, Bret H. Goodpaster, Kate Duchowny, Megan Hetherington-Rauth, Theresa Mau, Mahalakshmi Shankaran, Marc Hellerstein, William J. Evans, Steven R. Cummings

## Abstract

**Background:** Different measures to assess muscle size - magnetic resonance (MR) derived thigh muscle volume and d3-creatine dilution derived muscle mass (D_3_Cr muscle mass) – may have similar associations with strength, power, physical performance, fitness, and functional limitations in older adults.

**Methods:** Men (N=345) and women (N=482) aged ≥70 years from the Study of Muscle, Mobility and Aging completed exams including leg extension strength (1-repetition max) and cardiopulmonary exercise testing to assess fitness (VO_2_peak). Correlations and adjusted regression models stratified by sex were used to assess the association between muscle size measures and study outcomes; we tested for sex interactions.

**Results:** D_3_Cr muscle mass and MR thigh muscle volume were correlated (men: r=0.62, women: r=0.51, p<.001). Lower D_3_Cr muscle mass and lower MR thigh muscle volume were associated with lower strength and lower VO_2_peak in both men and women; D_3_Cr muscle mass was more strongly associated with strength in men than in women (p-int<0.05). There were correlations, though less consistent, between muscle size or mass with physical performance and function. Associations between the muscle size measures and the study outcomes occasionally varied by sex, and associations of MR thigh muscle volume were, at times, slightly more strongly associated with the study outcomes than was D_3_Cr muscle mass.

**Conclusions:** Less muscle –measured by either D_3_Cr muscle mass or MR thigh muscle volume - was associated with lower strength and worse performance. Varied associations by sex and assessment method suggest consideration be given to which measurement to use in future studies.

## Introduction

Muscle mass declines with age which is thought to correspond with reductions in strength, physical performance, self-reported function, and fitness.^1^ Many proxy and direct measures have been used to approximate muscle mass or volume, including those derived by bioimpedance analysis (BIA) or dual energy x-ray absorptiometry (DXA) for lean mass; computed tomography (CT) and magnetic resonance (MR) imaging for cross-sectional area or volume at a given anatomical site; and more recently, deuterated creatine (D_3_Cr) dilution for total body skeletal muscle mass. In general, measures of lean mass by DXA or BIA tend to be only modestly and inconsistently related to strength, performance and self-reported functional limitations.^2,3^ Measures of muscle size from other imaging techniques such as MRI or CT tend to have stronger associations with these outcomes,^2^ but a limitation to these imaging approaches is that that they only provide regional estimates, and, until recently, identification of muscle tissue required arduous manual segmentation. We and others have shown that low D_3_Cr muscle mass is consistently and strongly related to lower strength, lower performance, and greater functional limitations cross-sectionally and increased risk of disability, fractures and death over time; however, these data are largely limited to men.^4–7^ It remains unclear whether D_3_Cr muscle mass has similar relations to strength, function, and performance metrics in women as in men. Further, while D_3_Cr muscle mass is more consistently related to these outcomes than DXA derived measures of lean mass, it is not clear if D_3_Cr muscle mass and MR based measures of muscle size are similarly related to these strength, performance, and function measures.

Therefore, in the Study of Muscle, Mobility and Aging (SOMMA), a prospective multicenter cohort study of older adults, we aimed to quantify associations of muscle mass assessed with D_3_Cr and MR-based muscle volume with measures of strength and power; physical performance; fitness, and functional limitations, and to examine whether these associations differed by sex.

## Methods

### Study design

Participants aged 70 years and older were recruited to participate in SOMMA between April 2019 – December 2021 at two clinical centers (University of Pittsburgh and Wake Forest University School of Medicine). The detailed study design and description of measures in the SOMMA cohort are provided elsewhere.^8^ Briefly, participants must have been eligible for and willing to complete the muscle tissue biopsy and MR protocols; individuals were excluded if they 1) reported inability to walk ¼ mile or climb a flight of stairs; 2) had active cancer or with an advanced chronic disease such as heart failure, renal failure on dialysis, Parkinson’s disease, or dementia; 3) were unable to complete the 400-meter walk and walk faster than 0.6 m/s over 4 meters; and 4) had a BMI<40 kg/m^2^. The SOMMA Baseline Visit generally consisted of three days of assessments over several weeks: Day 1 (the 400-meter walk plus most other in-person assessments, 5 hours); Day 2 (cardiopulmonary exercise testing (CPET) and MR, 2-3 hours); and Day 3 (muscle biopsy, 2 hours); participants also completed a self-administered questionnaire. The protocol was reviewed and approved by the WIRB-Copernicus Group (WCG) IRB (20180764) and all participants gave informed consent.

### Assessment of body composition and size

Weight was measured using a balance beam or digital scales and height by wall-mounted stadiometers; body mass index (BMI) was calculated as weight (kg)/height (m^2^). An approximately 6-minute long MR scan was taken of the whole body to assess body composition including thigh muscle volume, muscle fat infiltration (% fat by proton density fat fraction), and adipose tissue volume in intra-abdominal and subcutaneous compartments (a proxy for total body fat) by AMRA Medical.^9^ Whole body D_3_Cr muscle mass was measured in participants using a standard protocol. Briefly, participants took a tablet with 30mg of D_3_-creatine and provided a fasting, morning urine sample 72-144 hours later.^10,11^ From the urine sample, D_3_-creatinine enrichment, unlabeled creatinine, and creatine were measured using high performance liquid chromatography-tandem mass spectroscopy; these measures were then included in a modified algorithm to determine total body creatine pool size and thus skeletal muscle mass as previously described.^11^ The modified algorithm for spillage correction, which accounts for potential loss of D_3_Cr dose, is as follows:

For Cr/Crn ratio <0.015, no spillage correction is applied

For Cr/Crn ratio 0.015-0.15, spillage correction (mg) = (exp((0.9424 × ln(Cr/Crn ratio)) - 0.1314)) × 30

For Cr/Crn ratio >0.15, spillage correction (mg) = (exp((1.6246 × ln(Cr/Crn ratio)) - 1.895)) × 30

Importantly, because the ratio of enrichments within the creatinine molecule is measured (i.e., the ratio of D_3_-creatinine to unlabeled creatinine), this method is not dependent on creatinine recovery, clearance or renal function. The method does not require any special dietary control other than the need for a fasting morning spot urine sample. In SOMMA, most participants had their D_3_-creatine dose and urine collection timed to the “Day 3” visit (muscle biopsy collection visit). This occurred by the clinic staff (at the “Day 1” or “Day 3” visit) determining the date for the dose administration from the scheduled date of the “Day 3” muscle biopsy collection visit, and then calculating the window for when the D_3_-creatine dose should be taken. Participants were then provided with the dose and instructions for when to take the dose at home, and staff made follow-up phone calls to remind participants about when to take the dose.

### Assessment of strength, power, physical performance, and functional limitations

Muscle strength and power were measured using the Jamar hand-held dynamometer (grip strength; Sammons Preston Rolyan, Bolingbrook, IL, USA)^12^ and the Keiser AIR300 or A420 Leg Press system (leg extension power and one repetition max, 1RM; Keiser Sports Health Equipment, Fresno, CA).^13^ Physical performance tests included a usual pace 400-meter walk; the Short Physical Performance Battery (SPPB)^14^ which included a timed 4-meter walk, chair stands, and balance test (side-by-side stance, modified tandem stance, and tandem stand); performance on each test was given a score of 0-4, and then summed. Those with an SPPB score ≤8 were considered to have an SPPB limitation.^15^ SOMMA men and women also completed the four-square step test,^16^ where participants were asked to step forward, backward and sideways into different quadrants to test balance; and a stair climbing task, where participants were asked to climb up and down four stairs three times without stopping.^17^ A balance test consisted of time held in 4 positions in this order: side-by-side stand (10 sec), semi-tandem stand (30 sec), tandem stand (30 sec), and one leg stand (30 sec). Total balance times ranged from 0-100 sec. Due to the skewed distribution, we analyzed the balance test times as a dichotomous variable (poor balance was defined as a total time <61 seconds). An expanded SPPB test was also administered that included the SPPB components plus a narrow walk (4 meters within a 20 cm course) and a single leg stand (max 30 seconds) that was computed as previously described with a range from 0-4.^18^ CPET on a treadmill using a modified Balke or manual protocol was used to estimate oxygen consumption at VO_2_peak.^19^

### Functional status

Functional status was assessed by questionnaire. Participants who reported any difficulty walking 2-3 blocks (1/4 mile) or climbing 10 steps were considered to have self-reported mobility limitation. We also classified people as having a functional limitation if they reported any difficulty with the following tasks: heavy housework, long distance walking (walking 1 mile), or difficulty getting out of bed or chairs. Functional status was also assessed with the Mobility Assessment Tool-short form (MAT-sf), which uses videos that depict a wooden mannequin performing a wide range of physical activities alongside a question about the participant’s self-reported ability to perform the task.^20–22^ ^20–22^ MAT-sf limitation was defined as a MAT-sf score of <60 (range 30-80).^23^

### Other variables

Age in years was based on reported date of birth; self-reported sex (man or woman) and race based on self-selection of current census categories; smoking, alcohol use and education level based on self-report. The Community Healthy Activities Model Program for Seniors (CHAMPS) questionnaire assessed specific types and the context of physical activities.^24,25^ Participants self-reported a physician diagnosis of arthritis, cancer (excluding non-melanoma skin cancer), cardiac arrhythmia, chronic kidney or renal disease, coronary heart disease, congestive heart failure, diabetes, osteoporosis, stroke, and aortic stenosis. Presence of depressive symptoms was determined by the Center for Epidemiologic Studies Depression Scale, short form (CESD-10; score of ≥10 indicated presence of depressive symptoms).^26^ We summed these self-reported conditions and presence of depressive symptoms to compute a modified version of the Rochester Epidemiology Studies multimorbidity index.^27^

### Statistical analysis

Given *a priori* knowledge of sex differences in body size, composition, strength, physical performance and functional limitations, all analyses were stratified by sex. We used t-tests, chi-square tests, and Wilcoxon rank-sum tests, as appropriate, for the nature and distribution of the variables, to compare characteristics of participants by sex. To determine the presence of a linear trend for characteristics across sex-specific tertiles of D_3_Cr muscle mass and MR thigh muscle volume, we used linear regression for normally distributed continuous variables, and two-sided Jonckheere-Terpstra test for trend for skewed and categorical variables. To quantify the relation between D_3_Cr muscle mass or MR thigh muscle volume with strength, power, and physical performance measures, we calculated correlation coefficients, unadjusted and adjusted for height and weight, and displayed associations as scatterplots. We considered correlations ≤|0.2| as weak, between |0.2| and |0.4| as moderate, and ≥|0.4| as strong. Logistic regression was used to separately estimate the likelihood of self-reported mobility limitation, functional limitation, MAT-sf limitation, poor balance, or SPPB limitation per standard deviation decrement in D_3_Cr muscle mass or MR thigh muscle volume. Models were adjusted for age, clinic site, weight, height, activity, and comorbidities. Similarly, we used adjusted linear regression models to estimate the association between D_3_Cr muscle mass or MR thigh muscle volume with the strength, performance and fitness measures. Finally, to determine whether differences in the association between muscle size (D_3_Cr muscle mass or MR thigh muscle volume) and our measures of strength, power, physical performance and functional limitation varied by sex, we tested an interaction term in models that included only the main effects and their product (that is, sex*muscle size). All analyses were completed in SAS 9.4 (Cary, NC).

## Results

Of the 879 SOMMA participants, 827 (94%) had data for D_3_Cr muscle mass and at least one outcome (Supplemental Figure 1). In SOMMA, men were more likely than women to report White race (Table 1). Men also reported greater alcohol intake, but smoking status, education level, and overall activity level did not differ between men and women. Women reported a greater number of chronic health conditions, including arthritis. Women were shorter and weighed less than men, although BMI and total abdominal adipose tissue did not significantly differ between men and women. Men had 44% greater D_3_Cr muscle mass (men: 26.8±6.1SD kg, women 18.6±4.1SD kg, p<.001), and 49% greater MR thigh muscle volume (men: 11.2±1.6SD L, women 7.5±1.1SD L, p<.001) than women. Women had greater muscle fat infiltration than men. Women were also weaker (lower grip strength, leg 1RM, and peak power) than men. Women had slower stair climb performance, but there were no significant differences between men and women in four square step performance or repeat chair stands. Men walked faster than women (400m walk, 4m walk, narrow walk) and had higher VO_2_peak. Men had borderline better performance on the expanded SPPB than women. Finally, men were higher functioning than women, based on the MAT-sf, self-reported mobility limitation, and functional limitation than women.

**Table 1.**
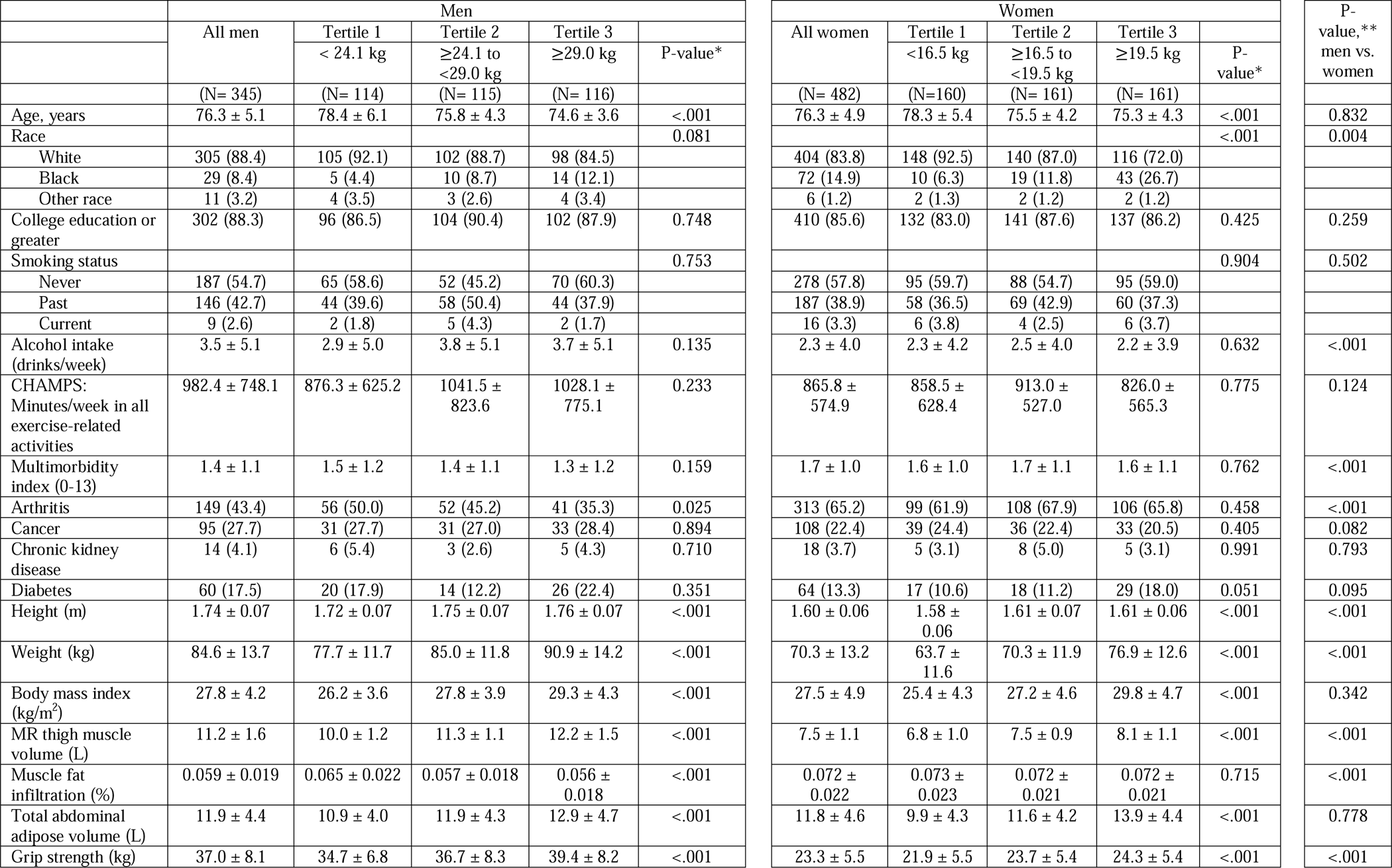
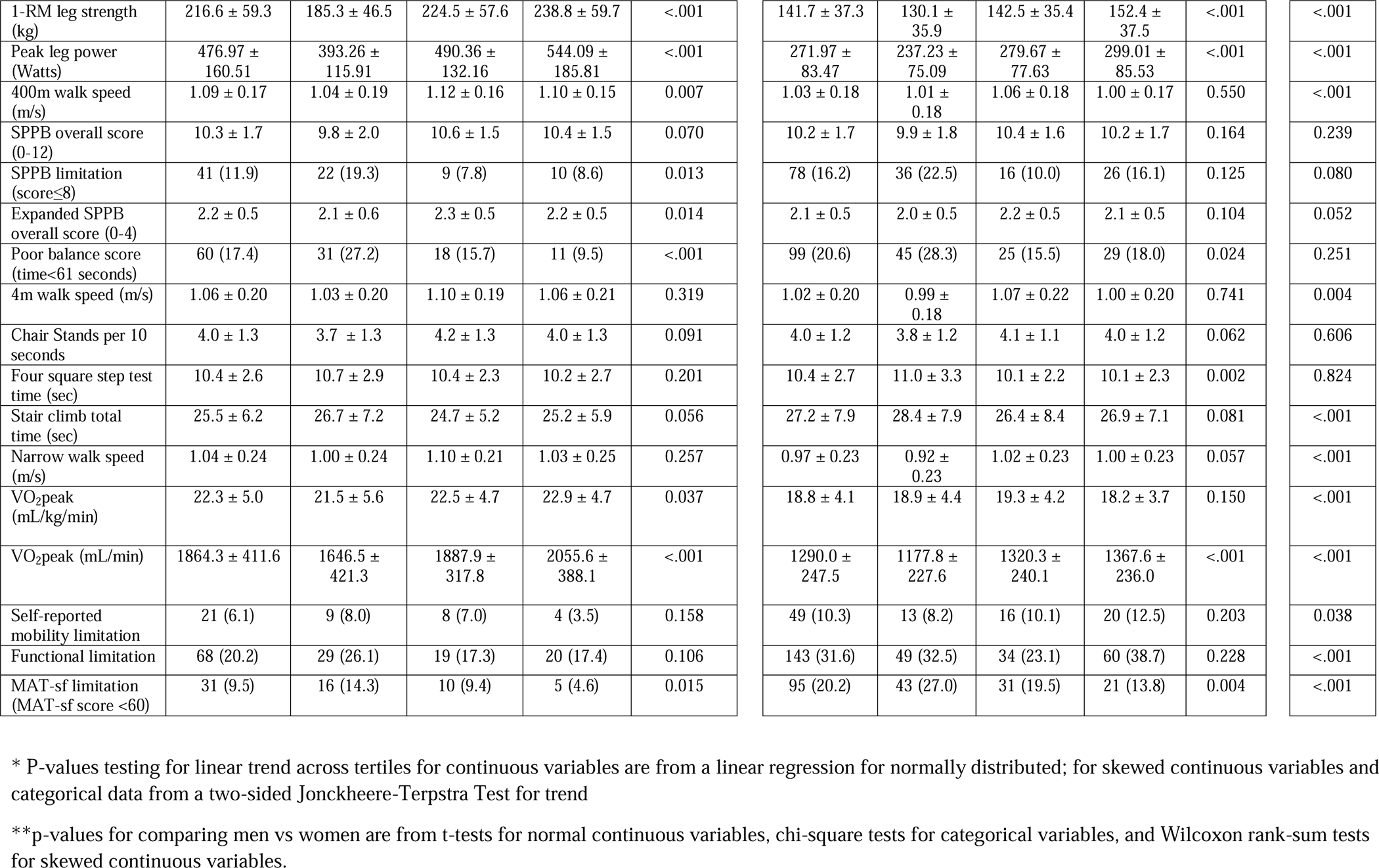
Characteristics of Men and Women, Overall and by Sex-Specific Tertile of D_3_Cr muscle mass in the Study of Muscle Mobility and Aging (SOMMA)

|  | Men |  |  |  |  | Women |  |  |  |  | P-value,**<br>men vs. women |
| --- | --- | --- | --- | --- | --- | --- | --- | --- | --- | --- | --- |
|  | All men | Tertile 1<br>< 24.1 kg | Tertile 2<br>≥24.1 to<br><29.0 kg | Tertile 3<br>≥29.0 kg | P-value* | All women | Tertile 1<br><16.5 kg | Tertile 2<br>≥16.5 to<br><19.5 kg | Tertile 3<br>≥19.5 kg | P-value* |  |
|  | (N= 345) | (N= 114) | (N= 115) | (N= 116) |  | (N= 482) | (N=160) | (N= 161) | (N= 161) |  |  |
| Age, years | 76.3 ± 5.1 | 78.4 ± 6.1 | 75.8 ± 4.3 | 74.6 ± 3.6 | <.001 | 76.3 ± 4.9 | 78.3 ± 5.4 | 75.5 ± 4.2 | 75.3 ± 4.3 | <.001 | 0.832 |
| Race |  |  |  |  | 0.081 |  |  |  |  | <.001 | 0.004 |
| White | 305 (88.4) | 105 (92.1) | 102 (88.7) | 98 (84.5) |  | 404 (83.8) | 148 (92.5) | 140 (87.0) | 116 (72.0) |  |  |
| Black | 29 (8.4) | 5 (4.4) | 10 (8.7) | 14 (12.1) |  | 72 (14.9) | 10 (6.3) | 19 (11.8) | 43 (26.7) |  |  |
| Other race | 11 (3.2) | 4 (3.5) | 3 (2.6) | 4 (3.4) |  | 6 (1.2) | 2 (1.3) | 2 (1.2) | 2 (1.2) |  |  |
| College education or greater | 302 (88.3) | 96 (86.5) | 104 (90.4) | 102 (87.9) | 0.748 | 410 (85.6) | 132 (83.0) | 141 (87.6) | 137 (86.2) | 0.425 | 0.259 |
| Smoking status |  |  |  |  | 0.753 |  |  |  |  | 0.904 | 0.502 |
| Never | 187 (54.7) | 65 (58.6) | 52 (45.2) | 70 (60.3) |  | 278 (57.8) | 95 (59.7) | 88 (54.7) | 95 (59.0) |  |  |
| Past | 146 (42.7) | 44 (39.6) | 58 (50.4) | 44 (37.9) |  | 187 (38.9) | 58 (36.5) | 69 (42.9) | 60 (37.3) |  |  |
| Current | 9 (2.6) | 2 (1.8) | 5 (4.3) | 2 (1.7) |  | 16 (3.3) | 6 (3.8) | 4 (2.5) | 6 (3.7) |  |  |
| Alcohol intake (drinks/week) | 3.5 ± 5.1 | 2.9 ± 5.0 | 3.8 ± 5.1 | 3.7 ± 5.1 | 0.135 | 2.3 ± 4.0 | 2.3 ± 4.2 | 2.5 ± 4.0 | 2.2 ± 3.9 | 0.632 | <.001 |
| CHAMPS: Minutes/week in all exercise-related activities | 982.4 ± 748.1 | 876.3 ± 625.2 | 1041.5 ± 823.6 | 1028.1 ± 775.1 | 0.233 | 865.8 ± 574.9 | 858.5 ± 628.4 | 913.0 ± 527.0 | 826.0 ± 565.3 | 0.775 | 0.124 |
| Multimorbidity index (0-13) | 1.4 ± 1.1 | 1.5 ± 1.2 | 1.4 ± 1.1 | 1.3 ± 1.2 | 0.159 | 1.7 ± 1.0 | 1.6 ± 1.0 | 1.7 ± 1.1 | 1.6 ± 1.1 | 0.762 | <.001 |
| Arthritis | 149 (43.4) | 56 (50.0) | 52 (45.2) | 41 (35.3) | 0.025 | 313 (65.2) | 99 (61.9) | 108 (67.9) | 106 (65.8) | 0.458 | <.001 |
| Cancer | 95 (27.7) | 31 (27.7) | 31 (27.0) | 33 (28.4) | 0.894 | 108 (22.4) | 39 (24.4) | 36 (22.4) | 33 (20.5) | 0.405 | 0.082 |
| Chronic kidney disease | 14 (4.1) | 6 (5.4) | 3 (2.6) | 5 (4.3) | 0.710 | 18 (3.7) | 5 (3.1) | 8 (5.0) | 5 (3.1) | 0.991 | 0.793 |
| Diabetes | 60 (17.5) | 20 (17.9) | 14 (12.2) | 26 (22.4) | 0.351 | 64 (13.3) | 17 (10.6) | 18 (11.2) | 29 (18.0) | 0.051 | 0.095 |
| Height (m) | 1.74 ± 0.07 | 1.72 ± 0.07 | 1.75 ± 0.07 | 1.76 ± 0.07 | <.001 | 1.60 ± 0.06 | 1.58 ± 0.06 | 1.61 ± 0.07 | 1.61 ± 0.06 | <.001 | <.001 |
| Weight (kg) | 84.6 ± 13.7 | 77.7 ± 11.7 | 85.0 ± 11.8 | 90.9 ± 14.2 | <.001 | 70.3 ± 13.2 | 63.7 ± 11.6 | 70.3 ± 11.9 | 76.9 ± 12.6 | <.001 | <.001 |
| Body mass index (kg/m<sup>2</sup>) | 27.8 ± 4.2 | 26.2 ± 3.6 | 27.8 ± 3.9 | 29.3 ± 4.3 | <.001 | 27.5 ± 4.9 | 25.4 ± 4.3 | 27.2 ± 4.6 | 29.8 ± 4.7 | <.001 | 0.342 |
| MR thigh muscle volume (L) | 11.2 ± 1.6 | 10.0 ± 1.2 | 11.3 ± 1.1 | 12.2 ± 1.5 | <.001 | 7.5 ± 1.1 | 6.8 ± 1.0 | 7.5 ± 0.9 | 8.1 ± 1.1 | <.001 | <.001 |
| Muscle fat infiltration (%) | 0.059 ± 0.019 | 0.065 ± 0.022 | 0.057 ± 0.018 | 0.056 ± 0.018 | <.001 | 0.072 ± 0.022 | 0.073 ± 0.023 | 0.072 ± 0.021 | 0.072 ± 0.021 | 0.715 | <.001 |
| Total abdominal adipose volume (L) | 11.9 ± 4.4 | 10.9 ± 4.0 | 11.9 ± 4.3 | 12.9 ± 4.7 | <.001 | 11.8 ± 4.6 | 9.9 ± 4.3 | 11.6 ± 4.2 | 13.9 ± 4.4 | <.001 | 0.778 |
| Grip strength (kg) | 37.0 ± 8.1 | 34.7 ± 6.8 | 36.7 ± 8.3 | 39.4 ± 8.2 | <.001 | 23.3 ± 5.5 | 21.9 ± 5.5 | 23.7 ± 5.4 | 24.3 ± 5.4 | <.001 | <.001 |
| 1-RM leg strength (kg) | 216.6 ± 59.3 | 185.3 ± 46.5 | 224.5 ± 57.6 | 238.8 ± 59.7 | <.001 | 141.7 ± 37.3 | 130.1 ± 35.9 | 142.5 ± 35.4 | 152.4 ± 37.5 | <.001 | <.001 |
| Peak leg power (Watts) | 476.97 ± 160.51 | 393.26 ± 115.91 | 490.36 ± 132.16 | 544.09 ± 185.81 | <.001 | 271.97 ± 83.47 | 237.23 ± 75.09 | 279.67 ± 77.63 | 299.01 ± 85.53 | <.001 | <.001 |
| 400m walk speed (m/s) | 1.09 ± 0.17 | 1.04 ± 0.19 | 1.12 ± 0.16 | 1.10 ± 0.15 | 0.007 | 1.03 ± 0.18 | 1.01 ± 0.18 | 1.06 ± 0.18 | 1.00 ± 0.17 | 0.550 | <.001 |
| SPPB overall score (0-12) | 10.3 ± 1.7 | 9.8 ± 2.0 | 10.6 ± 1.5 | 10.4 ± 1.5 | 0.070 | 10.2 ± 1.7 | 9.9 ± 1.8 | 10.4 ± 1.6 | 10.2 ± 1.7 | 0.164 | 0.239 |
| SPPB limitation (score≤8) | 41 (11.9) | 22 (19.3) | 9 (7.8) | 10 (8.6) | 0.013 | 78 (16.2) | 36 (22.5) | 16 (10.0) | 26 (16.1) | 0.125 | 0.080 |
| Expanded SPPB overall score (0-4) | 2.2 ± 0.5 | 2.1 ± 0.6 | 2.3 ± 0.5 | 2.2 ± 0.5 | 0.014 | 2.1 ± 0.5 | 2.0 ± 0.5 | 2.2 ± 0.5 | 2.1 ± 0.5 | 0.104 | 0.052 |
| Poor balance score (time<61 seconds) | 60 (17.4) | 31 (27.2) | 18 (15.7) | 11 (9.5) | <.001 | 99 (20.6) | 45 (28.3) | 25 (15.5) | 29 (18.0) | 0.024 | 0.251 |
| 4m walk speed (m/s) | 1.06 ± 0.20 | 1.03 ± 0.20 | 1.10 ± 0.19 | 1.06 ± 0.21 | 0.319 | 1.02 ± 0.20 | 0.99 ± 0.18 | 1.07 ± 0.22 | 1.00 ± 0.20 | 0.741 | 0.004 |
| Chair Stands per 10 seconds | 4.0 ± 1.3 | 3.7 ± 1.3 | 4.2 ± 1.3 | 4.0 ± 1.3 | 0.091 | 4.0 ± 1.2 | 3.8 ± 1.2 | 4.1 ± 1.1 | 4.0 ± 1.2 | 0.062 | 0.606 |
| Four square step test time (sec) | 10.4 ± 2.6 | 10.7 ± 2.9 | 10.4 ± 2.3 | 10.2 ± 2.7 | 0.201 | 10.4 ± 2.7 | 11.0 ± 3.3 | 10.1 ± 2.2 | 10.1 ± 2.3 | 0.002 | 0.824 |
| Stair climb total time (sec) | 25.5 ± 6.2 | 26.7 ± 7.2 | 24.7 ± 5.2 | 25.2 ± 5.9 | 0.056 | 27.2 ± 7.9 | 28.4 ± 7.9 | 26.4 ± 8.4 | 26.9 ± 7.1 | 0.081 | <.001 |
| Narrow walk speed (m/s) | 1.04 ± 0.24 | 1.00 ± 0.24 | 1.10 ± 0.21 | 1.03 ± 0.25 | 0.257 | 0.97 ± 0.23 | 0.92 ± 0.23 | 1.02 ± 0.23 | 1.00 ± 0.23 | 0.057 | <.001 |
| VO<sub>2</sub>peak (mL/kg/min) | 22.3 ± 5.0 | 21.5 ± 5.6 | 22.5 ± 4.7 | 22.9 ± 4.7 | 0.037 | 18.8 ± 4.1 | 18.9 ± 4.4 | 19.3 ± 4.2 | 18.2 ± 3.7 | 0.150 | <.001 |
| VO<sub>2</sub>peak (mL/min) | 1864.3 ± 411.6 | 1646.5 ± 421.3 | 1887.9 ± 317.8 | 2055.6 ± 388.1 | <.001 | 1290.0 ± 247.5 | 1177.8 ± 227.6 | 1320.3 ± 240.1 | 1367.6 ± 236.0 | <.001 | <.001 |
| Self-reported mobility limitation | 21 (6.1) | 9 (8.0) | 8 (7.0) | 4 (3.5) | 0.158 | 49 (10.3) | 13 (8.2) | 16 (10.1) | 20 (12.5) | 0.203 | 0.038 |
| Functional limitation | 68 (20.2) | 29 (26.1) | 19 (17.3) | 20 (17.4) | 0.106 | 143 (31.6) | 49 (32.5) | 34 (23.1) | 60 (38.7) | 0.228 | <.001 |
| MAT-sf limitation (MAT-sf score <60) | 31 (9.5) | 16 (14.3) | 10 (9.4) | 5 (4.6) | 0.015 | 95 (20.2) | 43 (27.0) | 31 (19.5) | 21 (13.8) | 0.004 | <.001 |

\* P-values testing for linear trend across tertiles for continuous variables are from a linear regression for normally distributed; for skewed continuous variables and categorical data from a two-sided Jonckheere-Terpstra Test for trend
\*\*p-values for comparing men vs women are from t-tests for normal continuous variables, chi-square tests for categorical variables, and Wilcoxon rank-sum tests for skewed continuous variables.

For both men and women, those with higher D_3_Cr muscle mass were younger and had larger body size and greater adiposity (i.e., height, weight, BMI, abdominal adiposity; Table 1), but there were no significant differences across D_3_Cr tertiles for physical activity, smoking, alcohol use, education, and number of multimorbidities. In women, but not in men, those with higher D_3_Cr were less likely to be White. For both men and women, those with higher D_3_Cr muscle mass were stronger (grip strength, 1-RM leg strength, peak leg power), had better balance, had higher fitness (VO_2_peak), and reported better function on the MAT-sf. Among men but not women, those with lower D_3_Cr muscle mass had worse performance on the expanded SPPB and 400m walk. Among women but not men, those with lower D_3_Cr muscle mass had worse performance on the four square step test. For men and women, there were no significant differences in SPPB overall score, chair stands, stair climb time, narrow walk, self-reported mobility limitations, and functional imitation by D_3_Cr muscle mass tertile, although a number of these associations were of borderline significance (0.05<p≤0.10) suggesting worse performance in lower D_3_Cr muscle mass tertiles. Characteristics of participants by tertiles of MR thigh muscle volume or D_3_Cr muscle mass/wgt by sex are shown in Supplemental Tables 1 and 2.

D_3_Cr muscle mass and MR thigh muscle volume were strongly correlated in both men and women (Figure 1, r=0.62 men, r=0.51 women, p<.001); accounting for body size (partial correlations adjusted for height and weight) reduced these correlations to moderate but they remained moderate or strong and statistically significant. In men, both D_3_Cr muscle mass and MR thigh muscle volume were positively correlated with leg strength, peak leg power (Figure 2) and grip strength (Supplemental Tables 3 and 4); correlations between these measures were generally smaller in women than in men (Table 2, p-interaction by sex for D_3_Cr muscle mass and grip strength=0.188; for leg strength=0.003; for leg peak power <.001; p-interaction by sex for MR thigh muscle volume and grip strength=0.492, for leg strength=0.139, and for leg peak power=0.007). Correlations between either D_3_Cr muscle mass or MR thigh muscle volume with 400m walk speed were weak (Figure 2) but were strengthened after adjusting for body size. MR thigh muscle volume was strongly correlated to VO_2_peak with and without adjustment for height and weight in both men and women; D_3_Cr muscle mass was strongly correlated with VO_2_peak in men and moderately correlated in women (Figure 2). In general, the point estimates for the correlation between MR thigh muscle volume and various strength and performance measures were higher than the point estimates between D_3_Cr muscle mass and the strength and performance measures; the confidence intervals of these estimates often excluded one another (Figure 2, Supplemental Tables 3 and 4) for both men and women.

**Figure 1.**
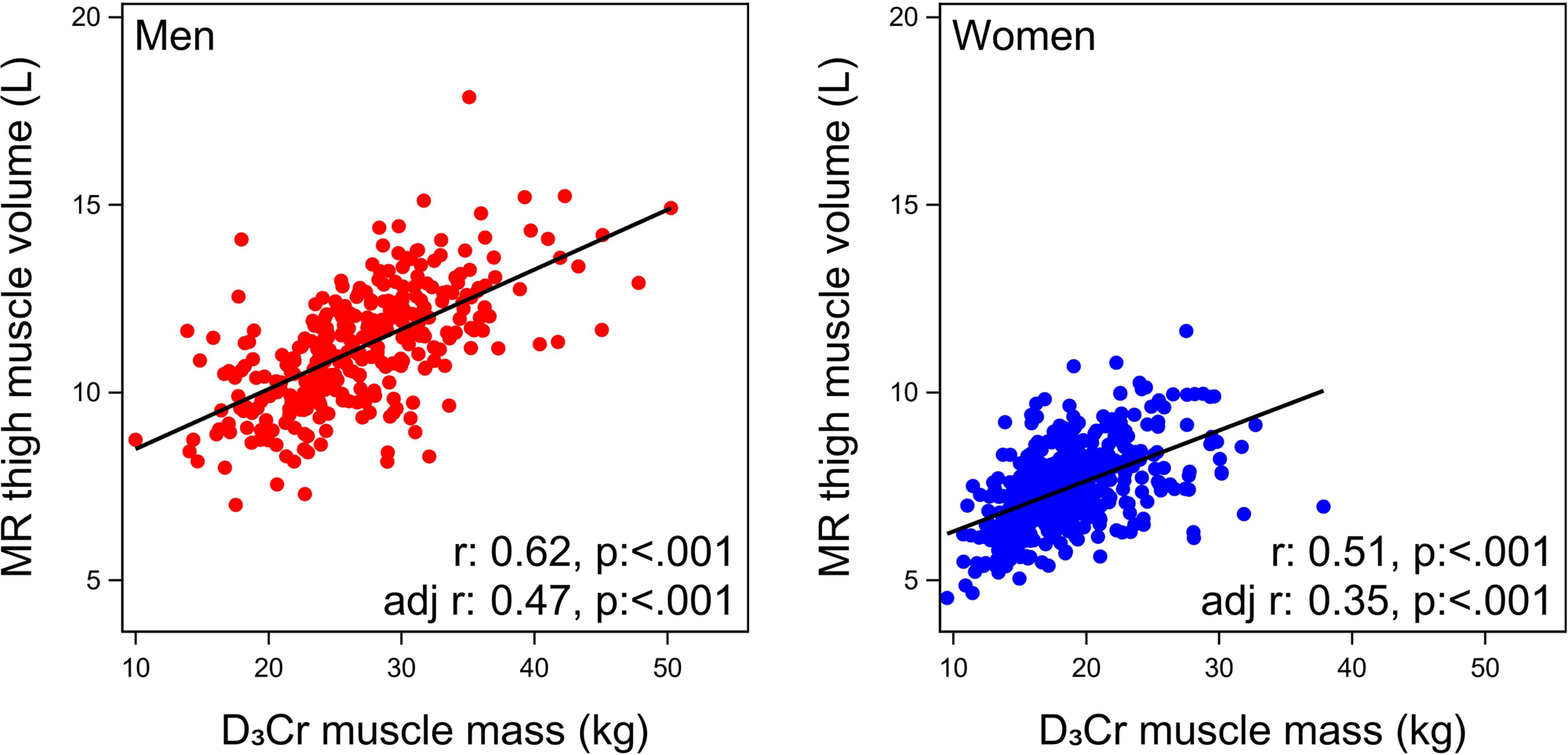
Pearson correlations (unadjusted, and adjusted for height and weight) between MR thigh muscle volume (L) vs. D_3_Cr muscle mass (kilograms) in men and women in SOMMA

**Figure 2.**
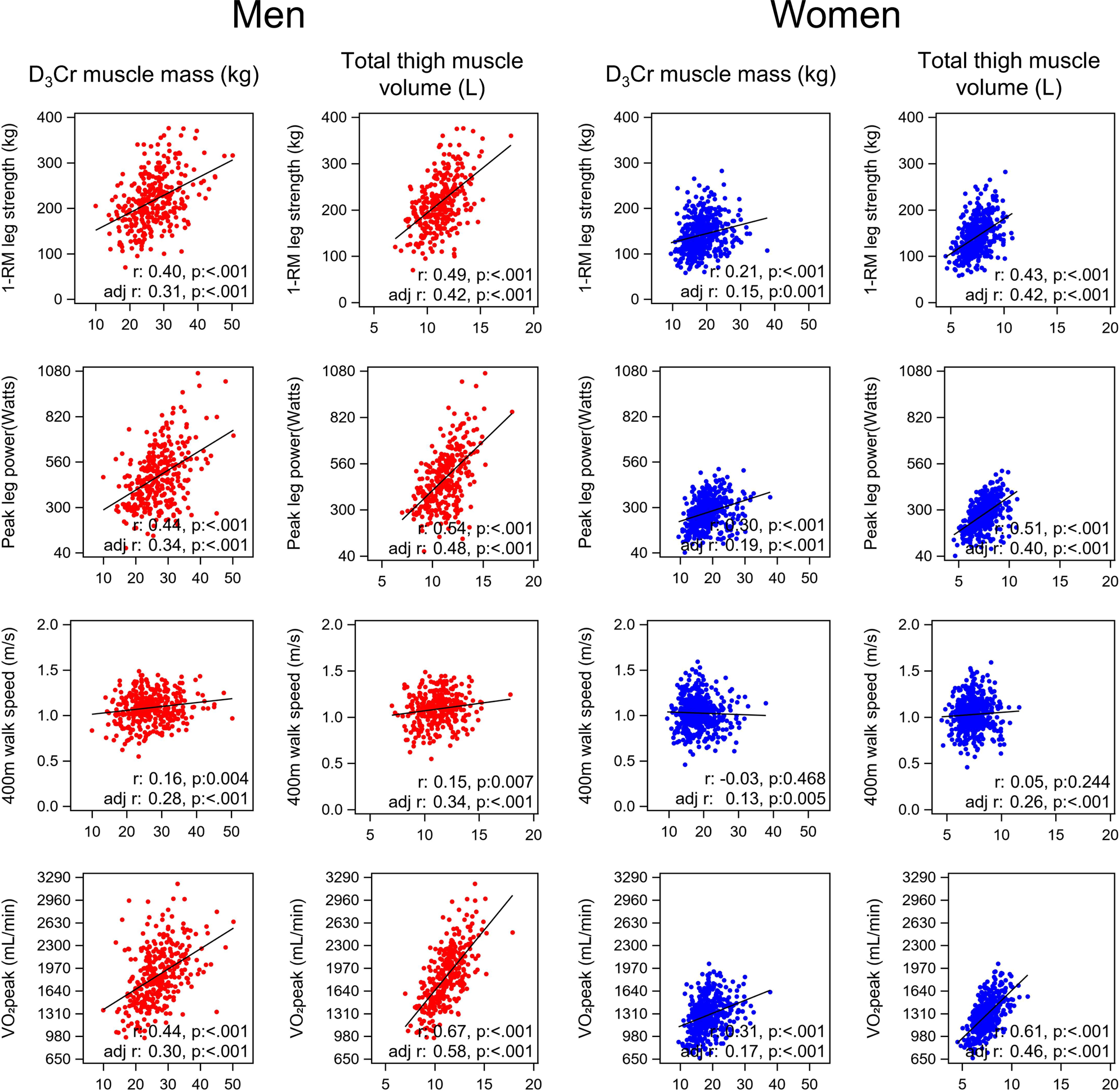
Pearson correlations (unadjusted and adjusted for height and weight) between D_3_Cr muscle mass and MR thigh muscle volume with 1-RM leg strength, peak leg power, 400m walk speed, and VO_2_peak in men (red) and women (blue) in SOMMA

**Table 2.** Multivariate adjusted association* of D_3_Cr muscle mass or MR thigh muscle volume with various strength and performance measures in older men and women (beta coefficient per sex-specific standard deviation decrement**).

|  | D<sub>3</sub>Cr muscle mass |  |  |  |  | MR thigh muscle volume |  |  |  |  |
| --- | --- | --- | --- | --- | --- | --- | --- | --- | --- | --- |
|  | Men |  | Women |  | p-int | Men |  | Women |  | p-int*** |
|  | N | Beta coefficient<br>(95% CI) | N | Beta coefficient<br>(95% CI) |  | N | Beta coefficient<br>(95% CI) | N | Beta coefficient<br>(95% CI) |  |
| Grip strength (kg) | 335 | **-1.1 (-2.0, -0.1)** | 472 | -0.3 (-0.8, 0.3) | 0.188 | 314 | **-2.5 (-3.7, -1.3)** | 449 | **-1.3 (-1.9, -0.6)** | 0.492 |
| 1-RM leg strength (kg) | 327 | **-14 (-21, -7)** | 458 | **-4 (-8, -1)** | 0.003 | 307 | **-32 (-40, -24)** | 435 | **-20 (-24, -16)** | 0.139 |
| Peak leg power (Watts) | 327 | **-44 (-61, -26)** | 458 | **-12 (-20, -5)** | <0.001 | 307 | **-89 (-110, -68)** | 435 | **-40 (-50, -31)** | 0.007 |
| 400m walk speed (m/s) | 337 | **-0.04 (-0.05, -0.02)** | 476 | -0.01 (-0.02, 0.00) | 0.019 | 316 | **-0.05 (-0.08, -0.03)** | 453 | **-0.04 (-0.06, -0.02)** | 0.449 |
| Expanded SPPB overall score (0-4) | 337 | **-0.11 (-0.17, -0.05)** | 474 | **-0.06 (-0.11, -0.02)** | 0.506 | 316 | **-0.16 (-0.23, -0.08)** | 451 | **-0.17 (-0.22, -0.11)** | 0.672 |
| 4m walk speed (m/s) | 337 | -0.02 (-0.05, 0.00) | 476 | -0.01 (-0.03, 0.01) | 0.446 | 316 | **-0.04 (-0.07, -0.01)** | 453 | **-0.04 (-0.06, -0.02)** | 0.582 |
| Chair stands per 10 seconds | 337 | **-0.24 (-0.39, -0.08)** | 475 | **-0.16 (-0.27, -0.05)** | 0.880 | 316 | **-0.42 (-0.62, -0.22)** | 452 | **-0.36 (-0.50, -0.22)** | 0.770 |
| Four square step test time (sec) | 308 | **0.35 (0.03, 0.68)** | 441 | **0.37 (0.10, 0.64)** | 0.406 | 291 | **0.59 (0.21, 0.97)** | 419 | **0.71 (0.38, 1.05)** | 0.242 |
| Stair climb total time (sec) | 328 | **1.27 (0.52, 2.01)** | 457 | **1.02 (0.33, 1.70)** | 0.957 | 307 | **1.43 (0.53, 2.33)** | 434 | **2.21 (1.35, 3.07)** | 0.482 |
| Narrow walk speed (m/s) | 303 | -0.01 (-0.04, 0.02) | 438 | **-0.02 (-0.05, -0.00)** | 0.558 | 285 | **-0.06 (-0.10, -0.02)** | 416 | **-0.05 (-0.08, -0.03)** | 0.493 |
| VO<sub>2</sub>peak (mL/min) | 322 | **-79 (-120, -38)** | 444 | **-30 (-50, -10)** | 0.016 | 304 | **-217 (-263, -171)** | 424 | **-111 (-135, -87)** | 0.010 |

\*models adjusted for age, site, weight, height, activity level, multimorbidity index
\*\*sex specific standard deviations: D<sub>3</sub>Cr muscle mass: men 6.13 kg, women 4.14 kg; MR thigh muscle volume: men: 1.58 L, women, 1.10 L
\*\*\* interaction models include muscle mass or volume, sex, and muscle mass or volume\*sex.
Beta coefficient (95% CI) shown in bold if p<0.05

After adjustment for age, clinic site, activity level, number of chronic medical conditions, and body size (height and weight), MR thigh muscle volume was related to all strength and performance measures in both men and women (Table 2). Of all measures examined, only the effect size for the associations of MR thigh muscle volume and leg power and VO_2_peak differed by sex (p-interaction<0.02). In contrast, the associations of D_3_Cr muscle mass with strength and performance differed by sex for a few measures. The associations of D_3_Cr muscle mass and leg power were stronger in men than women (p-interaction <0.01) as was the association with VO_2_peak (p-interaction=0.016). In men, there was a significant association of D_3_Cr muscle mass and grip strength and 400m walk speed, but no significant association among women.

Multivariable-adjusted associations of D_3_Cr muscle mass or MR thigh muscle volume with dichotomous measures of function were less consistent; this is perhaps due to the low prevalence of some limitations in SOMMA (e.g., only ∼6% of men and ∼10% of women self-reported a mobility limitation, Table 3) Men with lower levels of D_3_Cr muscle mass were more likely to have self-reported mobility limitation, functional limitation, MAT-sf limitation, SPPB limitation or poor balance, although the estimate for self-reported mobility limitation did not reach statistical significance. For women, lower D_3_Cr muscle mass was only significantly associated with MAT-sf limitation. We found evidence (p-int<0.05) that the association between D_3_Cr muscle mass and self-reported mobility limitation and functional limitation was stronger in men than it was in women. Similar to the findings with D_3_Cr muscle mass, men with lower MR thigh muscle volume were more likely to have self-reported mobility limitation, functional limitation, MAT-sf limitation, SPPB limitation or poor balance, although the estimates for MAT-sf limitation and SPPB limitation did not reach statistical significance. In contrast to the results for D_3_Cr muscle mass, in women, those with lower MR thigh muscle volume were more likely to have self-reported mobility limitation, functional limitation, MAT-sf limitation, SPPB limitation or poor balance, although the estimate for functional limitation did not reach statistical significance. There was no evidence that the relation between MR thigh muscle volume and the self-reported and objective measures of function varied by sex (p-int>0.05).

**Table 3.** Multivariate adjusted association* (odds ratio, 95% CI per sex-specific standard deviation (SD) decrement**) of D_3_Cr muscle mass or MR thigh muscle volume with dichotomous (self-reported and objective) measures of function in older men and women.

| Task or scale | D<sub>3</sub>Cr muscle mass |  |  |  |  | MR thigh muscle volume |  |  |  |  |
| --- | --- | --- | --- | --- | --- | --- | --- | --- | --- | --- |
|  | Men |  | Women |  | p-int*** | Men |  | Women |  | p-int*** |
|  | N of events (%) | Odds ratio (95% CI) | N of events (%) | Odds ratio (95% CI) |  | N of events (%) | Odds ratio (95% CI) | N of events (%) | Odds ratio (95% CI) |  |
| Self-reported mobility limitation | 21 (6.3) | 1.71 (0.94, 3.10) | 46 (9.8) | 0.96 (0.68, 1.36) | **0.061** | 20 (6.4) | **2.21 (1.05, 4.65)** | 44 (9.8) | **1.65 (1.01, 2.68)** | 0.547 |
| Functional limitation | 65 (20.1) | **1.68 (1.15, 2.46)** | 140 (31.3) | 0.89 (0.70, 1.13) | **0.021** | 57 (18.5) | **2.44 (1.48, 4.05)** | 136 (32.1) | 1.29 (0.94, 1.76) | 0.268 |
| MAT-sf limitation (score <60) | 31 (9.7) | **1.98 (1.16, 3.38)** | 93 (20.0) | **1.94 (1.41, 2.67)** | 0.936 | 26 (8.7) | 1.84 (0.93, 3.61) | 87 (19.7) | **2.56 (1.69, 3.87)** | 0.588 |
| SPPB limitation (score ≤8) | 41 (12.2) | **1.92 (1.20, 3.08)** | 75 (15.8) | 1.27 (0.94, 1.72) | 0.190 | 39 (12.3) | 1.49 (0.85, 2.60) | 71 (15.7) | **1.76 (1.17, 2.63)** | 0.978 |
| Poor balance (score <61 seconds) | 57 (20.4) | **2.32 (1.51, 3.55)** | 97 (20.4) | 1.21 (0.91, 1.59) | 0.342 | 56 (17.7) | **1.90 (1.17, 3.08)** | 95 (21.0) | **1.91 (1.32, 2.75)** | 0.357 |

\* Models adjusted for age, site, weight, height, activity level, multimorbidity index
\*\* D<sub>3</sub>Cr muscle mass SD: men 6.13 kg women 4.14 kg; MRI thigh muscle volume SD: men 1.58 L, women: 1.10 L.
\*\*\* interaction models include muscle mass or volume, sex, and muscle mass or volume\*sex.
Odds ratio (95% CI) shown in bold if p<0.05. N of events (%) is based on the total number of participants in each model.

## Discussion

We report several important findings. First, our results show that D_3_Cr muscle mass and MR thigh muscle volume are strongly related. Second, we found that muscle size (quantified by D_3_Cr muscle mass or MR thigh muscle volume) is strongly related to direct measures of strength and power in both men and women, but less strongly associated with more complex measures of physical performance. Third, in general, the associations between MR thigh muscle volume with strength, power, and physical performance were stronger than the associations of D_3_Cr muscle mass with these outcomes. Fourth, lower D_3_Cr muscle mass and lower MR thigh muscle volume had somewhat different associations with self-reported function outcomes, both in terms of agreement with each other and agreement between men and women.

D_3_Cr muscle mass and thigh MR muscle volume are related to one another, but, as expected, not perfectly correlated. The lack of perfect agreement is expected because they do not measure the same precise feature of skeletal muscle: MR is limited to estimates of fat-free of muscle volume in the thigh, while D_3_Cr muscle mass is a biochemical measurement of whole body creatine pool size from which total body muscle mass is derived.

Because sarcoplasmic creatine is found contiguous with contractile components,^28^ it may represent functional muscle. We do not have a full body measure of MR muscle volume, and we cannot regionalize measures of D_3_Cr muscle mass to specific anatomic regions. Each approach differs substantially in its methods, with different potential sources of error and variability; such different sources of error would have also lowered the correlation from 1.0. A complication in comparing the associations of the two methods to assess muscle size with our outcomes (i.e., strength, power, physical function, etc) is that the different anatomical regions (whole body vs thigh), measurement techniques and their errors and variability limit our ability to identify the precise reasons for any variations in the associations observed.

As expected, both low D_3_Cr muscle mass and low MR thigh muscle volume were related to lower strength in men and women. Such associations have been demonstrated previously with D_3_Cr muscle mass^5,29^ and MR based assessments of muscle size.^30,31^ The association of muscle size (MR thigh muscle volume or D_3_Cr muscle mass) with leg peak power and leg 1-RM was stronger in men than in women, but the p-for-interaction did not reach significance for the association of MR thigh muscle volume and 1-RM leg strength. It is somewhat surprising that the relationship between whole-body D_3_Cr muscle mass and leg strength is similar to that of thigh muscle volume. One interpretation of these findings is that whole-body D_3_Cr muscle mass reflects general, overall muscle function in older people. Muscle strength is a function of muscle size, motor recruitment patterns, and previous levels of physical activity.^32,33^ It is likely that in a relatively sedentary population, muscle mass is the strongest contributor to maximal force production.

When considering differences by sex for measures of performance and function, both low D_3_Cr muscle mass and low MR thigh muscle volume were less strongly and consistently related to measures of performance within and between sexes. In women and men, lower MR thigh muscle volume was similarly associated with all performance and fitness measures; only the association of MR thigh muscle volume with peak leg power was stronger in men than it was in women. In women and men, low D_3_Cr muscle mass was generally associated with worse performance with some exceptions. Our prior findings from the MrOS study parallel the associations we have observed here,^5,29^ but relatively little data have been previously published in women. One study of ∼80 women participating in the Women’s Health Initiative found strong associations between low D_3_Cr muscle mass and poor objective performance on the SPPB, and inconsistent associations with self-reported function.^7^ Taken together, our data suggests that muscle size may be a more important determinant of strength and power in men than it is in women, as has been suggested previously for other assessments of muscle size with strength or power.^34,35^ The sex differences in the association of muscle size with our study outcomes were more common for D_3_Cr muscle mass than for MR thigh muscle volume, suggesting that sex differences might depend to some extent on the measurement tool used.

These results may help clarify whether and how muscle size should be incorporated into clinical definitions of sarcopenia. There is a lack of consistent association of DXA based measures of lean mass with strength,^2,36^ physical performance, or self-reported function in men or in women. This had led some groups to propose that sarcopenia definitions omit estimates of muscle size based on DXA lean mass from diagnostic criteria, at least until a clinically feasible measure of muscle size that is related to functional outcomes is available.^36^ Our results indicate that both MR thigh muscle volume and D_3_Cr muscle mass are related to a variety of strength, function, performance and fitness measures in both men and women. Even with potential sex differences in these associations, our data suggest that MR thigh muscle volume or D_3_Cr muscle mass may be reasonable to include in clinical definitions of sarcopenia, especially if the associations with muscle function (i.e., strength and power) and other outcomes are verified with emerging data from studies of women, where data are particularly lacking. Exactly which measure to include in sarcopenia definitions (MR thigh muscle volume vs. D_3_Cr muscle mass) will depend on feasibility, reliability, and other issues related to clinical diagnostic tests. Future analyses using longitudinal data from both MR thigh muscle volume and D_3_Cr muscle mass will also provide addition insight into the relative utility of these measures, as loss of muscle mass or volume over time may be very different than static measures of low muscle mass or volume.

Our study has many strengths. SOMMA participants are extremely well-characterized with numerous assessments of strength, performance, fitness and self-reported function. However, we note some limitations. Our analysis is cross-sectional, and it is not clear if similar results would be seen if we examined associations of muscle size with changes in strength, performance, fitness and self-reported function. The SOMMA participants are highly selected, as they were free from mobility disability and dementia, without serious medical conditions, and eligible for our complex study measurements. Due to this study design, few participants at baseline were impaired or reported any limitations with function; exclusion of participants with low function might have underestimated associations of muscle mass or volume with strength, power, physical performance, fitness and functional limitations. Neither the MR thigh muscle volume measure nor the D_3_Cr muscle mass measure are yet clinically feasible for routine use. MR based measures exclude those with implanted metal and some devices, and those with claustrophobia. The D_3_Cr method is a timed test completed over several days that currently lacks regulatory clearance for clinical use. Thus, the immediate clinical impact of our results is limited. We did not adjust for multiple comparisons, so some of the reported significant results may have been due to chance. Finally, the SOMMA population is largely Non-Hispanic White, restricting our ability to complete our analyses within other race or ethnic subgroups; it is unclear whether these results generalize to other populations including younger adults.

In summary, our results demonstrate that muscle size (measured by either regional MR thigh muscle volume or by total body D_3_Cr muscle mass) is strongly related to strength and power in both men and women. Associations of muscle size with more complex measures of physical performance, fitness, and self-reported function outcomes is more variable, with potential differences for total body vs. regional measures and by sex. Varied associations by sex and assessment method suggest that consideration be taken when choosing the measurement of muscle size to use in future studies, which should focus on longitudinal associations with strength, power, physical performance, fitness, and functional limitations.

## Data Availability

All data produced are available online at sommaonline.ucsf.edu

https://sommaonline.ucsf.edu

## Acknowledgements

The Study of Muscle, Mobility and Aging is supported by funding from the National Institute on Aging, grant number AG059416. Study infrastructure support was funded in part by NIA Claude D. Pepper Older American Independence Centers at University of Pittsburgh (P30AG024827) and Wake Forest University (P30AG021332) and the Clinical and Translational Science Institutes, funded by the National Center for Advancing Translational Science, at Wake Forest University (UL1 0TR001420).

WJE and MKH are officers of MyoCorps, Inc a company that has licensed the granted patents for the D3creatine dilution method. PMC is a consultant and owns stock in Myocorps. SRC is a consultant to BioAge. No other authors report conflicts.

PMC drafted the manuscript and obtained funding. TLB completed the statistical analyses. SBK, ABN, RTH obtained funding and critically reviewed the manuscript. All other authors critically reviewed the manuscript.

